# THE REMOTE ANALYSIS OF BREATH SOUND IN COVID-19 PATIENTS: A SERIES OF CLINICAL CASES

**DOI:** 10.1101/2020.09.16.20195289

**Authors:** E. Furman, A. Charushin, E. Eirikh, G. Furman, V. Sokolovsky, S. Malinin, V. Sheludko

## Abstract

**Background:** Respiratory sounds have been recognized as a possible indicator of behavior and health. Computer analysis of these sounds can indicate of characteristic sound changes caused by COVID-19 and can be used for diagnosis of this illness.

**Purpose:** The communication aim is development of fast remote computer-assistance diagnosis of COVID-19, based on analysis of respiratory sounds.

**Materials and Methods:** Fast Fourier transform (FFT) was applied for analyses of respiratory sounds recorded near the mouth of 9 COVID-19 patients and 4 healthy volunteers. Sampling rate was 48 kHz.

**Results:** Comparing of FFT spectrums of the respiratory sounds of the patients and volunteers we proposed numerical healthy-ill criterions.

**Conclusions:** The proposed computer method, based on analysis of the FFT spectrums of respiratory sounds of the patients and volunteers, allows one to automatically diagnose COVID-19 with sufficiently high diagnostic values. This method can be applied at development of noninvasive self-testing kits for COVID-19.

## Introduction

WHO estimates that more than 18 000 000 people in the world are currently suffering from the novel coronavirus disease (COVID-19) [1]. COVID – 19 is a public health problem in countries regardless of their level of development. It is known that coronavirus SARS-CoV-2 causes severe lower respiratory disease with high mortality and evidence of systemic spread [2]. The virus is able to actively multiply in the epithelium of airways. Intense cough is one of the main symptoms of COVID-19 disease. It is known that the highest density of cough receptors is in the larynx [3]. Anatomically a dry cough can be associated with the effect of the virus on the cough receptors of the larynx due to infection with COVID-19. Coronavirus SARS-CoV-2 can penetrate into smallest airways, where it infects cells and causes bilateral pneumonia, and often with respiratory failure [4-7]. The defeat of various airways, caused by coronavirus, alters the sound formation of a patient and changes characteristics of respiratory sounds. Detection of the characteristic respiratory sounds (cough, wheezes, asthma wheezing, shortness of breath, etc.) is a widely used way of diagnostic of pulmonary diseases.

At present diagnosis of COVID-19 is based on clinical symptoms, and Chest X-ray / computer tomography, coronavirus tests (PCR (polymerase chain reaction) – molecular test, antigen test and specific antibodies to SARS CoV19) [6-8]. Due to very high contagiousness of COVID-19, development of screening diagnostic methods, including contactless and remote, is very relevant. One of these methods can be based on computer-assistance analysis of respiratory sounds of a patient and on comparison of the sound characteristics of a patient and a healthy volunteer.

The objectivity of auscultatory diagnostics can be significantly enhanced by using digitized audio signals and computer processing of these signals. The automated adventitious sound detection or/and their classification is a promising solution to overcome the limitations of conventional auscultation and to assist in the monitoring of relevant diseases, such as asthma, Chronic Obstructive Pulmonary Disease (COPD), and pneumonia [9]. Nemecio Olvera-Montes and co-authors [10] used the detection of respiratory crackle sounds via an android smartphone-based system for diagnostics of pneumonia and monitoring of the patient state.

Bersain A. Reyes and co-authors [11] used a smartphone-based system for automated bedside detection of crackle sounds in diffuse interstitial pneumonia patients. The performance of the automated detector was analyzed using: (1) synthetic fine and coarse crackle sounds randomly inserted to the basal respiratory sounds acquired from healthy subjects with different signal to noise ratios, and (2) real bedside acquired respiratory sounds from patients with interstitial diffuse pneumonia. In simulated scenarios, for fine crackles, an accuracy ranging from 84.86% to 89.16%, a sensitivity ranging from 93.45% to 97.65%, and a specificity ranging from 99.82% to 99.84% were found. The detection of coarse crackles was found to be a more challenging task in the simulated scenarios. In the case of real data, the results show the feasibility of using the developed mobile health system in clinical no controlled environment to help the expert in evaluating the pulmonary state of a subject.

The overview concern to the potential for computer audition (CA), i.e., the usage of speech and sound analysis by artificial intelligence to help in COVID-19 diagnostics [12]. The automatic recognition and monitoring of breathing, dry and wet coughing or sneezing sounds, speech under cold, eating behavior, sleepiness, or pain is used. The authors [12] concluded that CA appears ready for implementation of (pre-) diagnosis and monitoring tools.

It was consider that the acquired breathing sounds can be analyzed using advanced signal processing and analysis in tandem with new deep/machine learning and pattern recognition techniques to separate the breathing phases, estimate the lung volume, oxygenation, and to further classify the breathing data input into healthy or unhealthy cases [13]. Computer analysis of breath sounds can be able to important for identification of specific changes in these sounds, caused by COVID19.

Brown C. et co-authors use the exploring automatic diagnosis of COVID-19 from crowdsourced respiratory sound data [14]. The results of early works [11-14] and references therein allow one to suggest that breaths are useful sounds in COVID-19 diagnostics.

The **purpose** of the communication is development of a fast remote method of diagnosis of COVID-19 based on computer-assistance analysis of respiratory sounds. It is proposed to use a personal computer, modern mobile telephone, or smartphone for registration, recording, respiratory sounds and their analysis. The developed method can be applied as an additional method of COVID-19 diagnosis and as a personalized self-testing kits for COVID-19 virus. Such self-tests would serve as an early step before further procedures ordered by a medical doctor (lung CT, X-ray, etc.). The proposed method is based on analysis of Fast Fourier Transform spectrums and we restricted our work to use only breathing (and not cough and voice samples).

## Materials and Methods

The respiratory sounds were recorded by a smartphone Honor dua -1 22 at distance of 2 cm from the mouth for about seconds; sampling rate is 48 kHz. In the test 9 patients suffering from COVID-19 and 4 healthy volunteers have participated. COVID-19 in the patients has been diagnosed by standard medical methods. The clinical examination of the patients and the record of their respiratory sounds were made in Perm Infectious hospital and the volunteer sounds were recorded in the Perm State Medical University (Perm, Russia). The study complies with the declaration of Helsinki (adopted in June 1964, Helsinki, Finland) and revised in October 2000 (Edinburg, Scotland); the written agreements of the patients and volunteers have been obtained.

The beginning and end of the record contains temporal parts, in which respiratory sounds are not recorded, and “shots”. The shots can be much larger than a respiratory sound amplitude and do not reject the processes in airways. The applied preliminary processing removes these parts and also the highest shots

The proposed diagnosis method is based on that lung diseases cause changes in airways and these changes reject in spectrums of respiratory sounds. This approach has been applied in development of the computer-assistance methods of diagnostic of lung diseases [15-20]. The amplitudes of the harmonics of the Fast Fourier transform (FFT) are presented in Fig. 1. One can see several differences (Tabl. 1) in the spectrums for a volunteer and a patient, these differences can be used to formulate healthy-ill criterions, which are presented in Tabl. 2.

**Tabl. 1.** Comparison of the spectrums of a volunteer and a patient.

|  | Volunteer | Patient |
| --- | --- | --- |
| 1 | Minimum at about 2300 Hz | There is no extremum in the frequency range |
| 2 | Maximum at 3100 Hz | Minimum at 3300 Hz |
| 3 | Minimum at 4100 Hz | Maximum at 3900 Hz |
| 4 | Maximum at 4900 Hz | Minimum at 5000 Hz |
| 5 | There is no extremum at the frequency above 5300 Hz | Maximum at 5600 Hz |

**Tabl. 2.** Healthy-ill criterions.

Tabl. 2 Healthy-ill criterions
| Criteria | Healthy | Ill |
| --- | --- | --- |
| $k_1 = I(2300) / I(3200)$ | $k_1 < 1$ | $k_1 > 1$ |
| $k_2 = I(3200) / I(4000)$ | $k_2 > 1$ | $k_2 < 1$ |
| $k_3 = I(4000) / I(5000)$ | $k_3 < 1$ | $k_3 > 1$ |
| $k_4 = I(5000) / I(5600)$ | $k_4 > 1$ | $k_4 < 1$ |

**Fig. 1.**
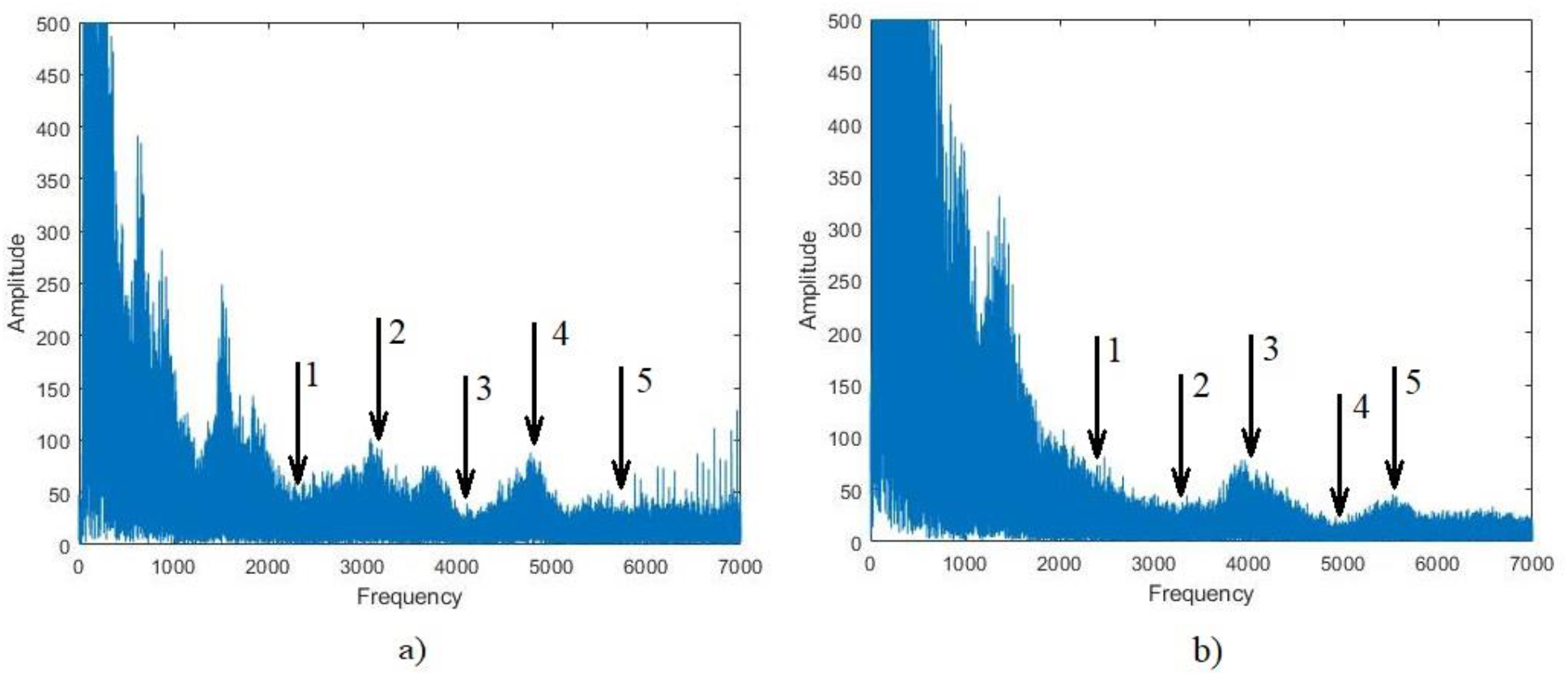
The amplitudes of the FFT harmonics for a healthy volunteer a) and a COVID-19 patient b). The amplitudes are given in arbitrary units.

In Tabl. 2

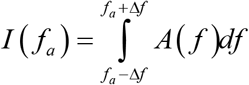

where *f*_*a*_ is the frequency of the extremum, *A*(*f*) is the harmonic amplitude at frequency *f*, Δ*f* is the frequency range, chosen equal 300 Hz. In program the integral is replaced by a sum of harmonic amplitudes with frequency in the range from *f*_*a*_ − Δ*f* to *f*_*a*_ − Δ*f*.

## Results

The results of the computer-assistance diagnostic for patients and volunteers are presented in Tabl. 3. In the table “x” notes coincidences of the computer-assistance and medical diagnoses; “0” notes the computer misdiagnosis.

**Tabl. 3.**
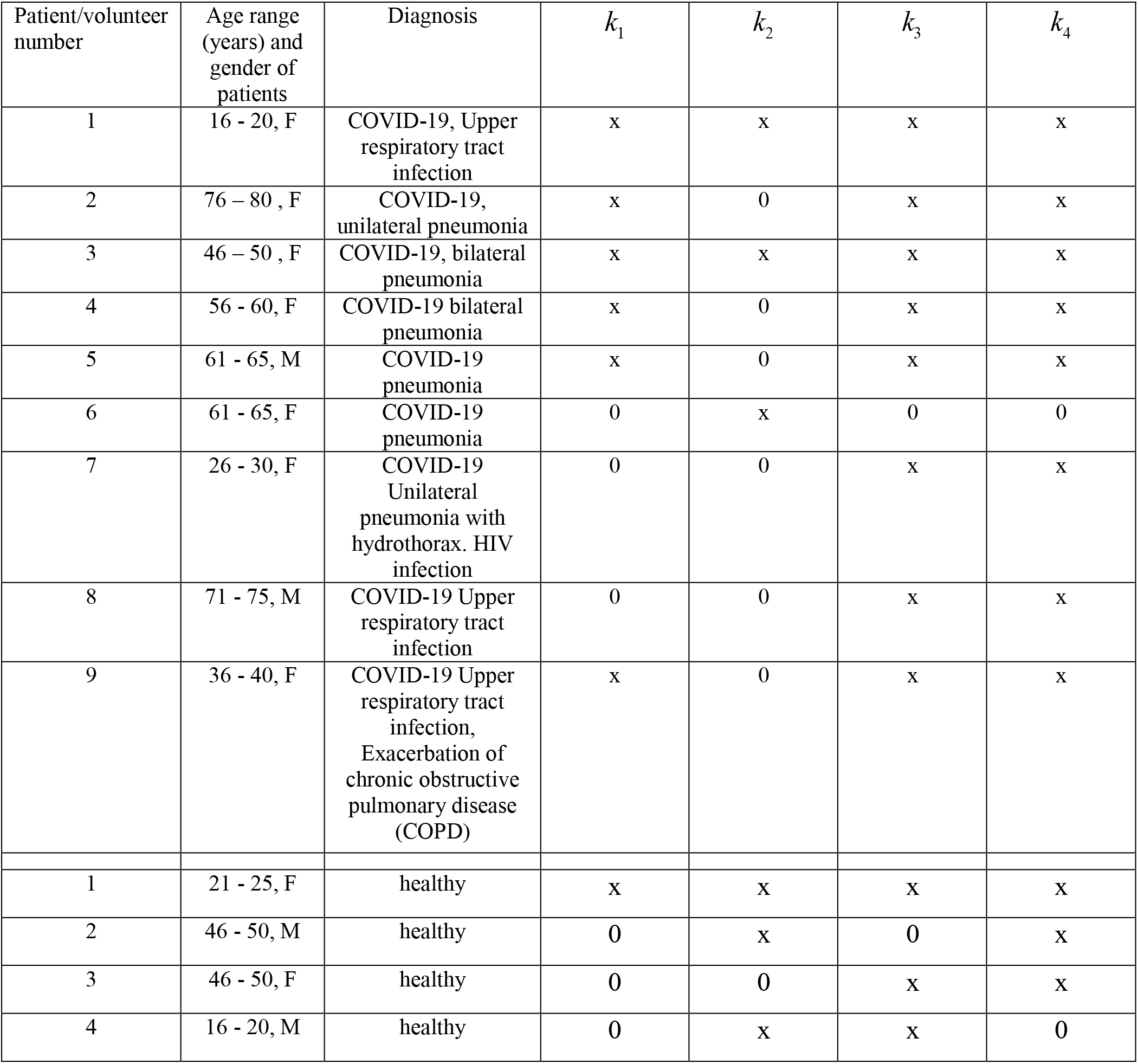
Results of the computer-assistance diagnostic for patients and volunteers.

| Patient/volunteer number | Age range (years) and gender of patients | Diagnosis | $k_1$ | $k_2$ | $k_3$ | $k_4$ |
| --- | --- | --- | --- | --- | --- | --- |
| 1 | 16 - 20, F | COVID-19, Upper respiratory tract infection | x | x | x | x |
| 2 | 76 - 80, F | COVID-19, unilateral pneumonia | x | 0 | x | x |
| 3 | 46 - 50, F | COVID-19, bilateral pneumonia | x | x | x | x |
| 4 | 56 - 60, F | COVID-19 bilateral pneumonia | x | 0 | x | x |
| 5 | 61 - 65, M | COVID-19 pneumonia | x | 0 | x | x |
| 6 | 61 - 65, F | COVID-19 pneumonia | 0 | x | 0 | 0 |
| 7 | 26 - 30, F | COVID-19 Unilateral pneumonia with hydrothorax. HIV infection | 0 | 0 | x | x |
| 8 | 71 - 75, M | COVID-19 Upper respiratory tract infection | 0 | 0 | x | x |
| 9 | 36 - 40, F | COVID-19 Upper respiratory tract infection, Exacerbation of chronic obstructive pulmonary disease (COPD) | x | 0 | x | x |
| 1 | 21 - 25, F | healthy | x | x | x | x |
| 2 | 46 - 50, M | healthy | 0 | x | 0 | x |
| 3 | 46 - 50, F | healthy | 0 | 0 | x | x |
| 4 | 16 - 20, M | healthy | 0 | x | x | 0 |

For both the patients and the volunteers diagnostic based on the criterions and gives much better results than these are given by diagnostic based on the criterions *k*_1_ and *k*_2_. This fact can be explained by the following. Changes of airway characteristics, caused by a disease, lead to the appearance of specific additional noises or/and changes in the respiratory sound spectrum.

## Discussion

Each disease is characterized by specific changes of the airways. For example, bronchial asthma is characterized by airway obstruction and inflammatory process, which covers all airways, from central to peripheral parts of tracheobronchial tree (small bronchi) [21]. Asthmatic changes in the lungs cause typical respiratory sounds with the main frequency in the low frequency ranges between 100 and 1000 Hz [22-24] and between 400 and 1600 [22, 25]. It can be assumed that а upper respiratory tract injury leads to appearance of changes in a higher frequency part of the spectrum. This assumption was confirmed by exam of the 5-th volunteer. For this volunteer the analysis showed that the criterions *k*_1_ and *k*_2_ corresponded to the healthy state but the criterions and - the ill one. Letter it was clarified that the volunteer suffered from early bronchitis. COVID-19 first affects а upper respiratory tract and them “propagates” into lungs and affects smaller airways [6]. The numerical values of the criterions *k*_1_ and *k*_2_ are obtained by analysis of the spectrum parts of relatively low frequencies. So, the incorrect diagnoses obtained using the criterions *k*_1_ and *k*_2_ (e.g. the 7-th patient) can be result that COVID-19 does not “propagate” deep in the lungs yet.

Another reason of the low accuracy of diagnostic based on the criterions *k*_1_ and *k*_2_ can be more higher sensitivity of the parameters *f*_*a*_ and Δ*f* at low frequencies to individual characteristics of patients (age, sex, wight, etc.) and also to fatigue and anxiety. One of the ways to increase sensitivity and reliability of the proposed computer-assistance method is to create big database and determine the parameters *f*_*a*_ and Δ*f* using machine learning technique. Additional criterions should be determined to differential diagnosis of COVID-19 from other respiratory diseases.

The proposal computer-assistance method can be applied as an additional screening fast remote method diagnostic of COVID-19. The method is based on the computer analysis of the FFT spectrum of respiratory sounds and demonstrates sufficiently high diagnostic values. The method can be basis to develop noninvasive self-testing kits for COVID-19 virus. To increase these values a big database of respiratory sounds of patients suffering from COVID-19 and volunteers should be created.

## Data Availability

All data will be provided upon request

## ACKNOWLEDGMENTS

This research was supported by a grant from the Ministry of Science & Technology (MOST), Israel & Russian Foundation (RFBR) (the joint research project № 19-515-06001). Authors thank Dr. Victor Meerovich and Dr. Alex Panich for help in development of computer analysis and fulfilled discussion of results.

